# A factorial randomised controlled trial to examine the potential effect of a text-message based intervention on reducing adolescent susceptibility to e-cigarette use: A study protocol

**DOI:** 10.1101/2024.04.23.24306261

**Authors:** Courtney Barnes, Heidi Turon, Sam McCrabb, Stephanie Mantach, Lisa Janssen, Megan Duffy, Daniel Groombridge, Rebecca Kate Hodder, David Meharg, Elly Robinson, Caitlin Bialek, Seaneen Wallace, Lucy Leigh, Luke Wolfenden

## Abstract

**Introduction:** Adolescent e-cigarette use, globally and within Australia, has increased in recent years. In response, public health agencies have called for the development of education and communication programmes targeting adolescents. Despite such recommendations, few rigorous evaluations of such interventions currently exist. The main objective of this study is to examine the potential effect of a text-message intervention targeting parents and adolescents on adolescent susceptibility to e-cigarette use (e.g. intentions towards using e-cigarettes). Secondary objectives are to: (1) examine the effect of the intervention on adolescent e-cigarette and combustible tobacco use; and (2) examine the acceptability of the intervention.

**Methods and analysis:** A randomised controlled trial employing a 2x2 factorial design will be conducted with parent-adolescent dyads (aged 12-15 years). Dyads will be randomly allocated to one of four arms: Arm 1 - a text-message intervention delivered to adolescents only; Arm 2 – a text-message intervention delivered to the parents of adolescents only; Arm 3 – a text-message intervention delivered to both the parents and adolescents; and Arm 4 - an information only control, consisting of an e-cigarette factsheet provided to parents only. Participant recruitment commenced in April 2023 with the aim to recruit 120 parent-adolescent dyads. Data collection to assess study outcomes will occur at baseline, 6-, 12- and 24-months post the commencement of the intervention. The primary endpoint will be 6-month follow-up. The primary outcome will be adolescent susceptibility to e-cigarette use, assessed using validated items. Analyses of trial outcomes will be undertaken under an intention-to-treat framework, with all participants included in the analysis in the group they were allocated.

**Ethics and dissemination:** Ethics approval has been obtained from the University of Newcastle Human Research Ethics Committee (H-2022-0340). Findings will be disseminated in peer-reviewed journals and at conferences.

**Trial registration:** Prospectively registered with Australia New Zealand Clinical Trials Registry (ACTRN12623000079640).

**STRENGTHS AND LIMITATIONS:**

- This study will employ a rigorous factorial randomised controlled trial design to test the effectiveness of a text-message based intervention;
- This study will contribute to the limited existing evidence base, with no randomised controlled trials aiming to prevent adolescent e-cigarette use currently published yet several underway;
- Selection bias may occur if adolescents who are highly susceptible to e-cigarette use refuse to participate in the study.

## INTRODUCTION

E-cigarettes deliver an aerosol by heating a liquid that users breathe in, which typically contains a range of chemicals, including those that add flavour, as well as nicotine in some devices. Adolescent e-cigarette use, globally and within Australia, has steadily increased in recent years, with e-cigarettes now the most heavily used nicotine-containing products amongst adolescents (1, 2). E-cigarettes were first introduced into the commercial market as a smoking cessation aid, with systematic review evidence indicating e-cigarettes show promise in increasing quit rates amongst adult smokers compared to traditional nicotine replacement therapy (3).

Nonetheless research indicates adolescents are now using e-cigarettes recreationally. For example, findings from one Australian survey reported that 14% of adolescents aged 14 to 17 years reported e-cigarette ever-use, almost half (48%) of which had never smoked combustible tobacco (4). Research examining the factors associated with adolescent e-cigarette use suggests that there is a range of determinants to adolescent e-cigarette use. A recent scoping review including 240 studies (5) identified a number of factors associated with adolescent e-cigarette use, including interpersonal (i.e. knowledge and attitudes (6)), societal (i.e. social norms, peer pressure (7)) and environmental (i.e. availability and exposure to marketing (8)).

A recent systematic review of global evidence concluded that serious adverse effects posed by the use of e-cigarettes include poisoning, burns and immediate toxicity through inhalation, including seizures. The review also found that consuming substances found within some e-cigarettes, such as the additive vitamin E acetate predominately found within cannabis-containing products, can result in acute lung injury known as EVALI (e-cigarette or vaping product use associated lung injury) (9). Additional meta-analyses of prospective cohort studies report e-cigarette use is associated with increased risk of future cigarette smoking, alcohol and marijuana use, as well as increased inflammatory responses and harmful effects on respiratory outcomes (10-13). Given the link between e-cigarette use in adolescents and subsequent initiation of combustible tobacco use (10), preventing adolescent e-cigarette use is a public health priority and a potentially potent strategy to further reduce future combustible tobacco-related cancer burden (14).

In addition to legislative action to prohibit the supply of e-cigarettes to adolescents, the World Health Organization and other (inter) national public health agencies recommend education and communication public health programmes (15-17). Text-message based interventions have proven to be an effective education-based approach to improving other adolescent health behaviours, including combustible tobacco use, nutrition, and physical activity (18, 19). These types of interventions target adolescents, but also often target parents as a way of influencing their child’s behaviour (19, 20). Formative evaluations (e.g., pre/post designs) of e-cigarette programmes indicate that text-message programmes may also be an effective approach to preventing adolescent e-cigarette uptake, influencing adolescent attitudes, and improving abstinence (21, 22).

Despite showing promise, text-message interventions targeting adolescent e-cigarette prevention are yet to be evaluated through rigorous trial designs, such as randomised controlled trials (RCT). For example, a recent Cochrane review examining the effectiveness of interventions to prevent or cease child and adolescent e-cigarette use failed to identify any published RCTs of such interventions (23). Promisingly, the review identified 15 ongoing studies with the aim to prevent adolescent e-cigarette use, however, no ongoing prevention studies included in the review were testing text-message based interventions

As such, employing a RCT design to test the effectiveness of a text-message based intervention aimed at reducing adolescent susceptibility to e-cigarettes, and thus preventing future use, would contribute substantially to a currently limited evidence base. Additionally, employing a factorial RCT design, which enable a robust method of comparing the effect of two or more interventions (or combinations thereof) directly in the one trial within an RCT preventing adolescent e-cigarette use will allow us to efficiently test multiple intervention targets (i.e., parents and/or adolescents) and draw comparisons across the approaches. Such an approach has the potential to contribute to the scant evidence base in an efficient and robust manner. Given parents as well as adolescents themselves are influential and recommended targets of prevention interventions, (5) the overall aim of this study is to examine the potential effect of a text-message program targeting parents and/or adolescents to prevent adolescent e-cigarette use, using a 2x2 factorial randomised controlled trial.

## OBJECTIVES

The specific objectives of this study are to: (1) examine the potential effect of a text-message based intervention targeting parents and adolescents on adolescent susceptibility to e-cigarette use; (2) examine the effect of the intervention on adolescent e-cigarette and combustible tobacco use; and (3) examine the acceptability of the intervention amongst adolescents and parents.

## METHODS AND ANALYSIS

### Ethics and registration

The research will be conducted and reported in accordance with the requirements of the Consolidated Standards of Reporting Trials (CONSORT). This protocol is reported according to the Statement and Standard Protocol Items: Recommendations for Interventional Trials (SPIRIT) (24). Ethics approval has been obtained from the University of Newcastle Human Research Ethics Committee (H-2022-0340) and the trial was registered prospectively with Australian New Zealand Clinical Trials Registry (ACTRN12623000079640).

### Trial design and setting

A RCT employing a 2x2 factorial design will be conducted with parent-adolescent dyads, consisting of adolescents aged 12-15 years. The trial consists of four arms: Arm one - a text-message based intervention delivered directly to adolescents only; Arm two – a text-message based intervention delivered directly to the parents of adolescents only; Arm three – a text-message based intervention delivered to both the parents and adolescents concurrently; and Arm four - an information only control. The trial will be conducted across Australia, which is geographically diverse, encompassing urban, rural, and remote areas.

### Eligibility criteria

Parent-adolescent dyads located within Australia will be recruited. To be eligible: (1) parents and adolescents must have sufficient English to engage with the intervention; and (2) adolescents must own or have exclusive access to a mobile phone. Adolescents will not be excluded if they report ever e-cigarette and/or cigarette use, however, they will be analysed separately. Dyads will be ineligible to participate if the adolescent is <12 or >15 years of age at the commencement of the study. For the purpose of this research, ‘parents’ will be defined as a biological mother or father, or legal guardian that has the authority to consent to the participation of a minor in a human research project and will be hereafter referred to as ‘parents’ within this protocol and supporting documents.

### Recruitment procedures

Parent-adolescent dyads will be recruited via multiple strategies, including social media advertisements, contacting parents that have participated in previous research trials conducted by the research group, through distribution (e.g., at meetings, events, or newsletters) or promotion (e.g., social media or websites) of this study by partners or relevant organisations. Recruitment commenced in March 2023. Potentially interested individuals will be encouraged to complete an online Expression of Interest (EOI) form housed on REDCap, (25) a secure data collection platform. The EOI will also contain a Participant Information Statement.

Within one week of a parent completing the EOI, an experienced research assistant will conduct a short telephone call with the parent to confirm eligibility, obtain verbal parent consent to participate in the study and collect adolescent contact information. Parents will also be encouraged to discuss the study with their child if they have not already done so, prior to the research team contacting the adolescent. Within this telephone call, eligible parents who provide consent to participate will complete baseline data collection via a short survey. Adolescents will then be contacted via text message or telephone call (as selected by their parent) by an experienced research assistant to confirm eligibility and obtain adolescent assent to participate in the study. A copy of the adolescent Participation Information Statement will be provided. Eligible and assenting adolescents will then complete baseline data collection during this process via a short survey.

Dyads where the parent is eligible and provides consent to participate but their adolescent either does not assent or is ineligible will be ineligible for this study.

### Randomisation and blinding

Following baseline data collection, an independent statistician will randomise parent-adolescent dyads following a block randomisation procedure (using block sizes of 4 and 8) to one of the four trial arms (described below) using statistical analysis software (SAS 9.3) in a 1:1:1:1 ratio. Due to the geographic and socioeconomic diversity of Australia, randomisation will be stratified by dyad geographic location (rural/urban) and socio-economic status, as determined by 2016 Socio-Economic Indexes for Areas (SEIFA) categorisation (26) using dyad postcodes. Dyads ranked in the top 50% of Australia will be classified as least disadvantaged and the lower 50% of postcodes as most disadvantaged by the SEIFA categorisation. Dyads are unable to change arms following randomisation and allocation.

Due to the nature of the intervention, parent-adolescent dyads will be aware of their allocation. Data collection staff will not be informed of group allocation until after the collection of primary trial outcomes. Data analysis will be conducted by a statistician blinded to group allocation.

### Intervention

The text messages used within the intervention have been developed to target factors (i.e., barriers and enablers) associated with adolescent e-cigarette use identified through an extensive search of existing literature and a recent scoping review incorporating 240 studies (5). Findings of the scoping review identified interpersonal (e.g. e-cigarette knowledge, beliefs and attitudes; refusal skills) and social (e.g. influence of peers and family; availability of social support) factors as the most frequently reported modifiable factors associated with adolescent e-cigarette use (6-8). All identified factors were mapped to the domains of the Theory of Triadic Influence, a theory previously employed to guide the development of text-message based interventions to prevent and address adolescent combustible tobacco use (27). The theory consists of three domains: biology and personality (e.g. individual knowledge, propensity for risk taking behaviour, use of other substances); social context (e.g. influence of peers and parents); and broader environment (e.g. legislation, exposure to advertising) (27). The Behaviour Change Wheel was used to determine which identified factors could be targeted effectively within a text-message based intervention (28). Following the process outlined by Michie et al, factors were categorised using the COM-B model as either capability, opportunity or motivation (28). Behavioural change techniques were then selected to embed within the text messages in order to address the factors associated with adolescent e-cigarette use (Figure 1) (28). The content of the messages were developed following a co-design process with parents, adolescents, parenting research experts, tobacco and e-cigarette experts, and behavioural scientists. This iterative process included semi-structured interviews, online surveys, and text-message writing activities to develop and refine the messages.

**Figure 1.**
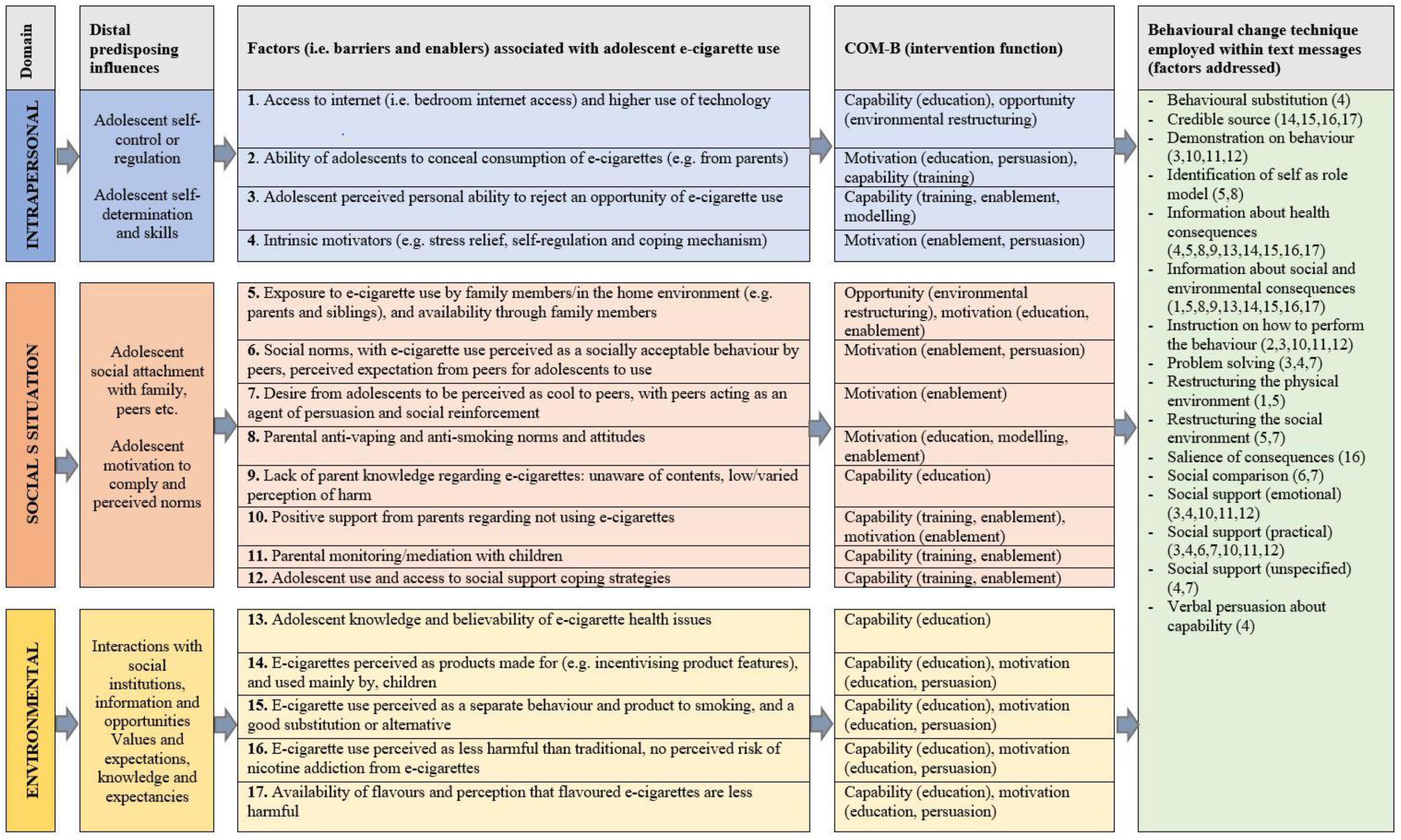
Logic model of text-message intervention.

The co-design process resulted in a bank of messages developed to address a range of factors associated with adolescent e-cigarette use (Figure 1). Each text-message targets one or more identified factors and embeds behavioural change techniques to prevent e-cigarette use. Specific text-messages will contain links to external evidence-based resources, including videos, factsheets and animations, which align with the factor(s) addressed within each text-messages (29-33). The content and timing of text-messages delivered to parents and adolescents (depending on arm allocation) will align, designed to foster healthy conversation amongst families.

### Arms

Parent-adolescent dyads will be randomly allocated to one of the following four arms:

#### Adolescent text-messages (Arm one)

Adolescents from the parent-adolescent dyads allocated to Arm one will receive a theoretically informed text-message intervention to prevent adolescent e-cigarette use. Messages will be sent to adolescents to target factors associated with adolescent e-cigarette use, aligned to each of the Theory of Triadic Influence domains (24) and embedding behavioural change techniques (25). These factors include knowledge regarding health effects, social norms and peer influence, and adolescent refusal skills (6-8). Adolescents will receive one message per week for 12 weeks.

Parents from the parent-adolescent dyads allocated to Arm one will not receive parent text-messages; however, they will receive an evidence-based e-cigarette factsheet developed by NSW Health to educate parents on the risks associated with child and adolescent e-cigarette use (26).

#### Parent text-messages (Arm two)

Parents from the parent-adolescent dyads allocated to Arm two will receive a theoretically informed text-message intervention to prevent adolescent e-cigarette use. Messages will be sent to parents to target factors associated with adolescent e-cigarette use that could be influenced by parents, such as adolescent accessibility and exposure to e-cigarettes, role of parents as a positive support mechanism, parent knowledge and perceptions of harms regarding adolescent e-cigarette (6-8). Like the adolescent messages, the parent messages align to each of the Theory of Triadic Influence domains (24) and embed behavioural change techniques (25). Parents will receive one message per week for 12 weeks and will also receive the e-cigarette factsheet described above (26).

Adolescents from the parent-adolescent dyads allocated to Arm two will not receive the adolescent text-messages nor any other information from the research team during the intervention period.

#### Parent and adolescent text-messages (Arm three)

Parent-adolescent dyads randomly allocated to Arm three will receive both the parent and adolescent text-messages described above. Parents and adolescents will receive the text-messages concurrently. Parents will also receive the e-cigarette factsheet described above (26).

#### Control (Arm four)

Parents from the parent-adolescent dyads allocated to the control arm will receive the e-cigarette factsheet described above only (26).

Distribution of the text-messages will be controlled centrally by the research team, limiting the potential for contamination between arms.

### Outcome measures

Data collection to assess study outcomes will occur at baseline and three follow-up time points: 6-, 12- and 24-months post the commencement of the intervention. Parents will only be invited to participate in data collection at baseline and 6-month follow-up, whereas adolescents will be invited to participate at all four time points. To reduce the potential for participant drop out or attrition, the study team will employ a range of participant retention strategies based on a recent Cochrane systematic review by Edwards and colleagues (34). A summary of study outcomes and data collection time points is provided in Figure 2.

**Figure 2.**
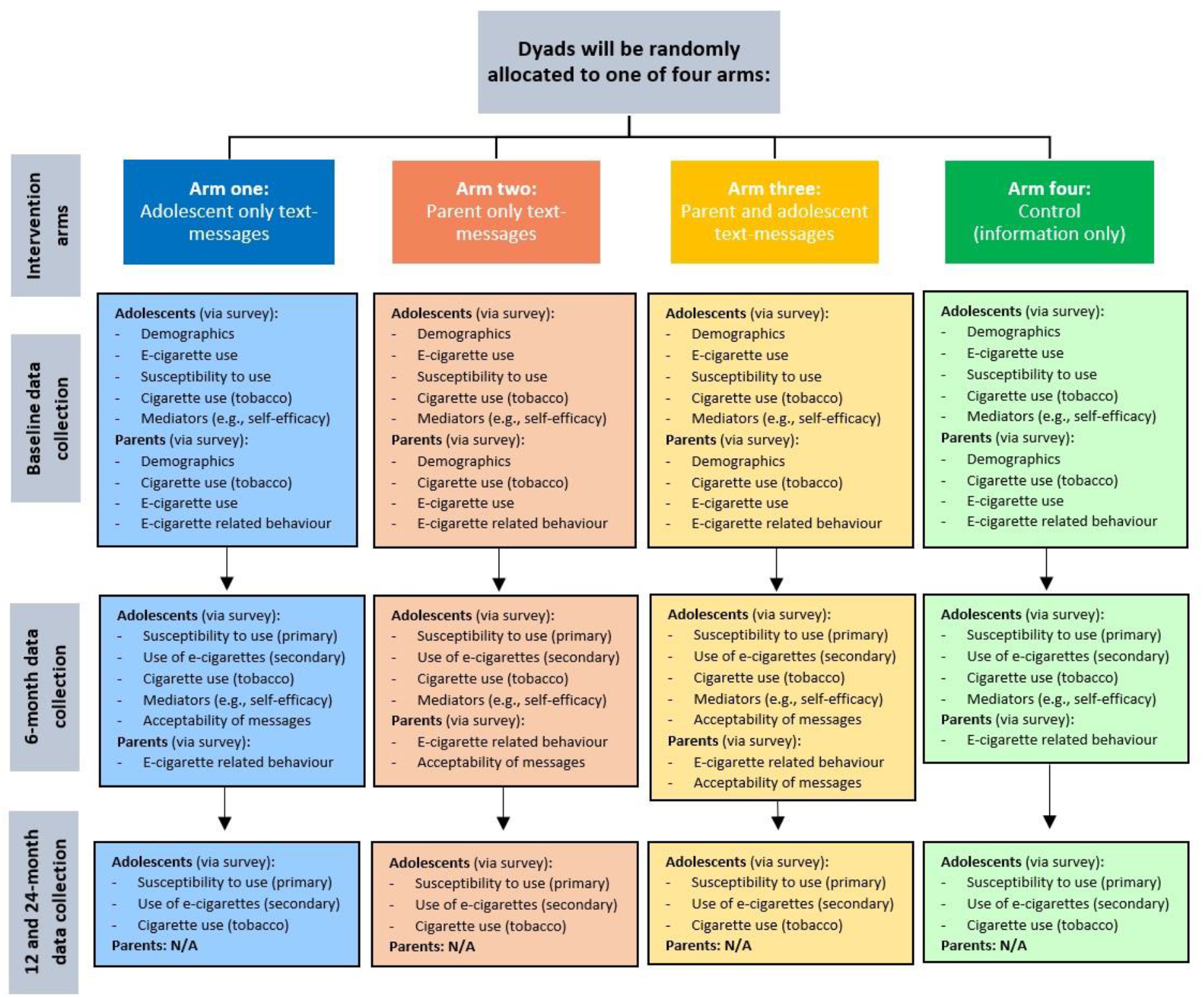
Study outcomes and data collection time points.

### Primary outcome

Adolescent susceptibility to e-cigarette use will be assessed in an online or telephone survey conducted by a trained research assistant at baseline, 6-, 12- and 24-month follow-up with adolescents only. The primary end point will be 6-month follow-up. Three validated survey items outlined by Carey et al. (35) will be used to assess adolescent susceptibility to use e-cigarettes. Adolescents will be asked to respond to each item on a 5-point scale, ranging from 1 (definitely not) to 5 (definitely yes). Adolescents will be categorised as non-susceptible to each individual item if they responded “Not at all curious” or “Definitely not,” with any other response categorised as susceptible (35). A mean susceptibility score will be calculated by summing scores across the three survey items (36).

### Secondary outcomes

Adolescent ever-use of e-cigarettes, defined as any lifetime use of e-cigarettes, will be assessed via an online or telephone survey conducted by a trained research assistant at baseline, 6-, 12- and 24-month follow-up with adolescents only (37).

Adolescent combustible tobacco-use will be assessed via an online or telephone survey conducted by a trained research assistant at baseline, 6-, 12- and 24-month follow-up with adolescents only. As recommended by the World Health Organization to measure youth tobacco use (37), survey items include ever-use of cigarettes (defined as lifetime use, including one or two puffs) and the current use (defined as use in the past 30 days) of other forms of tobacco, including cigars, pipes and smokeless tobacco.

#### Adolescent e-cigarette and combustible tobacco use behaviours

To aid comparisons of the effect of e-cigarette policies and interventions, Pearson et al. (38) recommends eight core constructs of e-cigarette use to be assessed. As such, adolescents that report e-cigarette ever-use at follow-up will be asked additional items within the survey to assess the remaining constructs, including: frequency of e-cigarette use; former daily use; relative perceived harm; device type; primary flavour preference; presence of nicotine within the e-cigarettes; and primary reason for use. Adolescents that report ever-use of cigarettes will be asked additional items including age of cigarette smoking initiation and current use of cigarettes (defined as use in the past 30 days).

#### Acceptability of the text-message intervention

Acceptability (defined as the perception amongst adolescents and parents that the intervention is agreeable, palatable or satisfactory (39)) will be assessed in an online or telephone survey at 6 months follow-up with adolescents (i.e. Arm one and Arm three) and parents (i.e. Arm two and Arm three) who received the text-messages. Survey items to assess acceptability will be based on validated items developed by Weiner et al. (40) and those previously used by the research team (41). Parents and adolescents will be asked to respond on a 5-point Likert scale, ranging from 1 (strongly agree) to 5 (strongly disagree).

#### Intervention mediators

Changes in the three Theory of Triadic Influence domains targeted by the intervention (i.e., adolescent self-efficacy and behavioural control; social normative beliefs; and knowledge and attitudes towards the behaviour) (24) that may work to drive changes in the primary and secondary outcomes will be assessed via survey with adolescents at baseline and 6-month follow-up. Validated survey items developed by Barker et al. (42) and modified items from a validated survey by Zourbanos et al. (43) have been mapped to the three Theory of Triadic Influence domains targeted by the intervention to determine how the intervention may influence e-cigarette intentions and behaviour. Adolescents will be asked to respond to each item on a 5-point scale, ranging from 1 (definitely wouldn’t) to 5 (definitely would).

Parent knowledge, behaviour and skills relating to e-cigarettes will be assessed via an online or telephone survey conducted by a trained research assistant at baseline and 6-month follow-up with parents in all study arms. Validated items from previous surveys assessing parental attitudes and behaviours towards other health behaviours (e.g. combustible tobacco smoking, alcohol use), will be modified to capture information on e-cigarette use and accessibility in the home environment, as well as parent knowledge of e-cigarettes and perceived confidence to discuss and provide support to adolescents regarding e-cigarettes (44-46). Parents will be asked to respond to each item on a 5-point scale, ranging from 1 (strongly agree) to 5 (strongly disagree).

### Participant Characteristics

#### Adolescent and parent characteristics

Adolescent and parent characteristics, including gender, age, location (to enable socioeconomic status and rural/urban status to be determined), cultural and linguistic diversity and self-reported Aboriginal or Torres Strait Islander status will be collected via an online or telephone survey at baseline.

### Process Measures

Parent and adolescent engagement with the text message program will be assessed using individual-level analytics automatically captured via the online text-message platform throughout the intervention period, including delivery rates and clicks on links to additional information within the text messages. Additional information regarding parent and adolescent recall of, and engagement with, the text-messages will be collected via the online or telephone survey with parents and adolescents at 6-months follow-up.

#### Intervention fidelity

Fidelity of the intervention, defined as the degree to which the intervention was implemented as it was prescribed in the original protocol or as it was intended by the program developers (i.e. the research team), (39) will be assessed through project records maintained by the research team and analytics from the text message program. Analytics will be used to determine the number of parents and adolescents in each arm (described above) that received each text message as intended and the timing of delivery.

#### Cultural evaluation

A cultural evaluation will also be conducted to explore the landscape and cultural implications of vaping within the local Aboriginal community. The cultural evaluation will also seek feedback from Aboriginal participants regarding the opportunities to embed, amend or strengthen the cultural responsiveness of the text-messages.

### Statistical analysis plan

#### Sample size

This trial will be undertaken in two phases (described below), with the first phase being used to pilot the intervention. The target sample size for phase one will be no less than 120 parent-adolescent dyads (30 dyads per group). Only dyads with consenting parents and adolescents will be included in the target sample size (i.e. parent only consent/participation will not count towards the target sample size). A formal sample size calculation has not been calculated for the piloting phase (phase 1) consistent with recommendations for pilot and feasibility studies (47). However, planned apriori analysis of data from the piloting phase will be used to submit a formal sample calculation for the fully powered trial following the recruitment and follow-up of the initial 120 dyads.

#### Analyses

Statistical analyses will be conducted using STATA v14. Descriptive statistics, including proportions, means and standard deviations, will be used to describe adolescent and parent characteristics, engagement with the intervention and acceptability of the text-message intervention.

An intention to treat approach will be used in the analyses of all trial outcomes, with all participants included in the analysis in the group that they were allocated. For the primary analyses, to assess the effects of the parent and adolescent message interventions separately, mixed effects regression models will be used to assess between group differences in strategies (parent messages vs no parent messages; and adolescent messages vs no adolescent messages) in adolescent outcomes at follow-up. For continuous outcomes (i.e. adolescent susceptibility to e-cigarette use), a linear mixed model will be used. For the dichotomized outcomes, a generalized linear mixed model with a binomial distribution and logit link function will be used. Each model will include a fixed effect for each intervention group. An additional fixed effect for the baseline value of the outcome will also be included in all linear mixed models. We will employ multiple imputation following ITT principles for any missing data. All statistical tests will be two-tailed with alpha of 0.05. We will examine the potential interaction between intervention strategies by graphically displaying outcome measures by group given the small sample size and the limited power to detect an interaction effect.

#### Data management

The participant information statement informs participants about the planned or possible future use of information/data. All hard copy information will be stored at the workplace of the research team at Hunter New England Population Health’s secure Wallsend location in locked filing cabinets and secure computer files. Only research personnel and approved staff working with the data will have access to the data. Any electronic data will only be accessible via password protected accounts and any file-sharing will be restricted to members of the project team. All identifying information will be kept for the required 7 years until it needs to be destroyed.

In the interest of open science we would like to be able to share de-identified data and provide access to any potential systematic reviews at the individual level as well as any required secondary analysis. Other researchers may seek access to the data for the purposes of re-analysis and secondary analysis. Any use of data that is not covered by the current ethics approval will require additional ethics approval before the data is made available. Anyone seeking to access the data will need to contact the lead investigator, along with seeking appropriate ethical clearances. Only once those approvals are granted will de-identified data be shared via an encrypted communication channel.

### Patient and public involvement

Parents and adolescents were involved in the development of the text message content following a co-design process with parents, adolescents, parenting research experts, tobacco and e-cigarette experts, and behavioural scientists. This iterative process included semi-structured interviews, online surveys, and text-message writing activities to develop and refine the messages. A multi-disciplinary advisory group, consisting of health promotion practitioners, behavioural scientists, Aboriginal health professionals and program managers, parenting researchers, tobacco and e-cigarette experts, will oversee the conduct of the trial.

## DISCUSSION

Public health interventions are recommended by national and international agencies to supplement policy and legislative approaches to address the emerging issue of adolescent e-cigarette use. Despite such recommendations, no randomised controlled trials to test the potential impact of a text-message based intervention on preventing adolescent e-cigarette use have been reported to date. Such interventions represent a potentially effective and rapidly scalable approach to address this public health crisis. This study, employing a rigorous trial design and developed to address proven determinants of adolescent e-cigarette use reported in the current literature, will contribute to a limited yet rapidly emerging evidence base.

## Data Availability

No data is reported within the manuscript.

## LIST OF ABBREVIATIONS

EOI: Expression of interest;
EVALI: e-cigarette or vaping product use associated lung injury;
ITT: Intention to treat;
CONSORT: Consolidated Standards of Reporting Trials;
RCT: Randomised controlled trial;
SEIFA: Socio-Economic Indexes for Areas (SEIFA);
SPIRIT: Statement and Standard Protocol
Items: Recommendations for Interventional Trials.

## DECLARATIONS

### Ethics and dissemination

Ethical approval has been provided by the University of Newcastle Human Research Ethics Committee (approval H-2022-0340). The trial is prospectively registered with the Australian New Zealand Clinical Trials Registry (ACTRN12623000079640). Consent from parents and adolescents to participate in the study with approved information statements and consent processes. Participants will provide either written or verbal consent to participate depending on the format they select to complete the eligibility and consent process (i.e. via telephone with a trained research assistant will collect verbal consent; online will collect written consent). Participants may withdraw from the study at any time. Evaluation and process data collected as part of the study will be disseminated widely through national and international peer-reviewed publications and conferences presentations.

## Consent for publication

Not applicable

## Availability of data and materials

Not applicable

## Authors’ contributions

First author Courtney Barnes led the development of this manuscript. Courtney Barnes, Caitlin Bialek, Sam McCrabb, Heidi Turon, Stephanie Mantach, Lisa Janssen, Megan Duffy and Luke Wolfenden led the development of the intervention, research design and evaluation. Lucy Leigh specifically revised the statistical analyses sections of this paper. Daniel Groombridge, Rebecca Kate Hodder, David Meharg, Elly Robinson and Seaneen Wallace provided guidance and feedback on the development of the study protocol. All authors contributed to the drafting and final approval of the manuscript.

### Trial oversight

A multi-disciplinary advisory group has been established and will oversee the conduct of the trial to ensure the research is co-produced. The advisory group consists of health promotion practitioners, behavioural scientists, Aboriginal health professionals and program managers, parenting researchers, tobacco and e-cigarette experts. A Terms of Reference has been developed for the group. The project team, consisting of research staff and health promotion practitioners, will conduct the study to align with the study protocol. Aboriginal advisory group members will provide cultural governance and guidance throughout the project. The advisory group will contribute to the dissemination of the study, including any publications or reports that are produced.

